# Using a Novel-Liaison Based Social Media Intervention Utilizing Community Feedback to Improve Organizational Health Literacy in Covid-19 Crisis Communication

**DOI:** 10.1101/2025.02.14.25322261

**Authors:** Nishita Matangi, Cristie Granillo, Maud Joachim-Célestin, Karla Estudillo Fuentes, Crissy Irani, Nikhil Thiruvengadam, N Susanne Montgomery

## Abstract

The COVID-19 pandemic exposed historical pervasive gaps in information and resources for vulnerable communities. In an academic community partnered response of a consortium of already collaborating community and academic partners, the COVID Care Corps (CCC), was developed to create and disseminate public health information to individuals in the Inland Empire (IE) by including community liaisons to implement a systematic feedback loop to assure community voice. Liaisons made disseminating information and getting community feedback more efficiently in a time of crisis when information needed to be immediate. As a result, it was found that although community was concerned about the pandemic, they also wanted information about other social determinants of health (immigration, finances, housing and job security= often presented as competing priorities to the pandemic. This resulted in a shift in content creation to aligned with community needs and interests. Establishing trust through community relationships and identifying community liaisons should be implemented prior to a crisis to ensure the quick dissemination of information needed for local buy-in and true equitable health communication.

## Introduction

The COVID-19 pandemic had a global impact and affected the lives of people worldwide; however, its effects were not felt equally. Within the United States, certain geographical areas were more impacted than others. These regions had higher poverty rates, multigenerational households and high minority populations (Athavale et al., 2021, Lundberg et al., 2020, Parra et al., 2022), which were often already disenfranchised and at higher risk for health disparities and bore a higher burden from the pandemic (Chowkwanyun & Reed, 2020, Nana-Sinkam et al., 2021). Being “essential workers” - they were further impacted by mandated measures requiring them to continue working (Nayak et al., 2020).

One such region is the Inland Empire in Southern California, which is made up of San Bernardino and Riverside Counties It’s one of the fastest growing areas in California, with more than half of the population identifying as African American, Latino, and Native American (Public Policy Institute of California, 2018). Among its residents, 63 percent of deaths come from cardiovascular diseases, respiratory conditions, cancer and diabetes (Prevention Institute, 2017), which are all conditions positively associated with a higher risk of mortality and serious complications from COVID-19 (Centers for Disease Control and Prevention, 2024).

As is the case with every epidemic, effective crisis communication from the start would have been vital to contain the COVID-19 pandemic. However, due to the novelty of the disease and the nature and process of communication shared, many people were overwhelmed and unable to differentiate between facts and opinions (Mohammed, et al., 2021). This resulted in a state of uncertainty translated into palpable fear and distrust. Healthy People 2030 differentiated organizational health literacy from personal health literacy shifting the responsibility of effective communication to organizations and challenging them to provide health information that is easy to find, understand and use to address community fears and concerns (Healthy People 2030). Assuming that it is accessible, having accurate information plays a vital role in communities’ ability to adopt healthy habits and follow guidelines that help its residents to protect themselves. This is especially true in emergency situations, like a pandemic. In addition, studies show that seeing accurate information prior to misinformation makes individuals less susceptible to believing the misinformation.

The concept of a community feedback loop and including community in the process of creating health information is not a new one. Several studies have shown that including community voice in the process is effective to reach the same community because it improves relevance and accessibility (Jackson et al., 2023, Adebisi & Rabe, 2021). Who those community voices are and how they are included is what is most important. When timely information is needed for a population that is at high risk for feeling the worst of crisis and emergencies, it can be easier to focus on just getting the information out, especially since it can take more effort and time to gather the feedback. But using it to guide communication can build trust and improve response to the communication (Yumiya et al., 2020)

Recognizing the need for clear and relevant information at the community level, one of the largest academic medical centers serving the Inland Empire quickly moved to develop an academic-community partnership committee comprised of partners from local community-based organizations, religious organizations, and school districts. The goal was to gather and prepare timely COVID-19 information that was scientifically accurate yet accessible and relevant to the Inland Empire’s diverse community. ***The purpose of this manuscript is to share the process, results and lessons learned in implementing this initiative that was designed to mitigate the communication gap during the first year of the COVID-19 pandemic through a collaborative partnership: the COVID Care Corps (CCC)*.**

## Materials and Methods

This retrospective study describes the bi-directional communication methods of CCC to improve organizational health literacy. By inviting ongoing community feedbackdissemination of public health information intentionally made to be more relatable and accessible to the community of focus. The main audiences for this project were under-resourced communities and families for which English is not the primary language spoken at home.

### Collaboration

This partnership was intended to create communication pathways between those who created the information and those who received the information through community liaisons. CCC recognized that feedback was important, but in the crisis setting of the pandemic it was necessary to find efficient ways to get that feedback without any type of bottleneck of information flow from either side. The figure below shows the different players and their roles creating and disseminating information and then sharing feedback to inform creation and dissemination of new information and this was done through previously established partnerships with community. The initial program began with the establishment of teams in early April of 2020, closely following the California stay-at-home order.

The academic side included a team made up of university faculty, staff and students who filled the roles of content developers, researchers and designers. When a topic was chosen, researchers pulled and then summarized educational and resource information. Then content developers outlined the content specifically for handouts or social media posts. Designers then designed the content and made it ready for dissemination which was then sent to community liaisons.

Community liaisons were individuals at each community organization that made up the community component of the partnership. There was direct contact with each of them and in most cases, they were the ones who were on the ground disseminating the information to their organization’s target population. If they were not directly disseminating the information they were directly in communication with those who were.

Community partners included school districts, churches, community organizations, community programs and even the Mexican consulate. Each of the organizations served populations in two of Californias largest counties. All of these population already faced significant health disparities and were disenfranchised with high rates of poverty, low education and literacy rates and many were either monolingual in a language other than English or spoke English as a second language. Many being in service industries and in multi-family and multi-generational homes were at higher risk for being exposed to and spreading Covid-19.

### The “feedback loop” process

Two teams were created to generate a system to develop, evaluate, and disseminate information to the community (see Figure 1). The first team was responsible for research and content development. – They outlined, prioritized, edited or translated information from reputable sources (e.g., the Centers for Disease Control and Prevention, local public health departments) to make them more accessible to the local community, and designed content for dissemination in different mediums including email, social media, and physical flyers.

**Figure 1.**
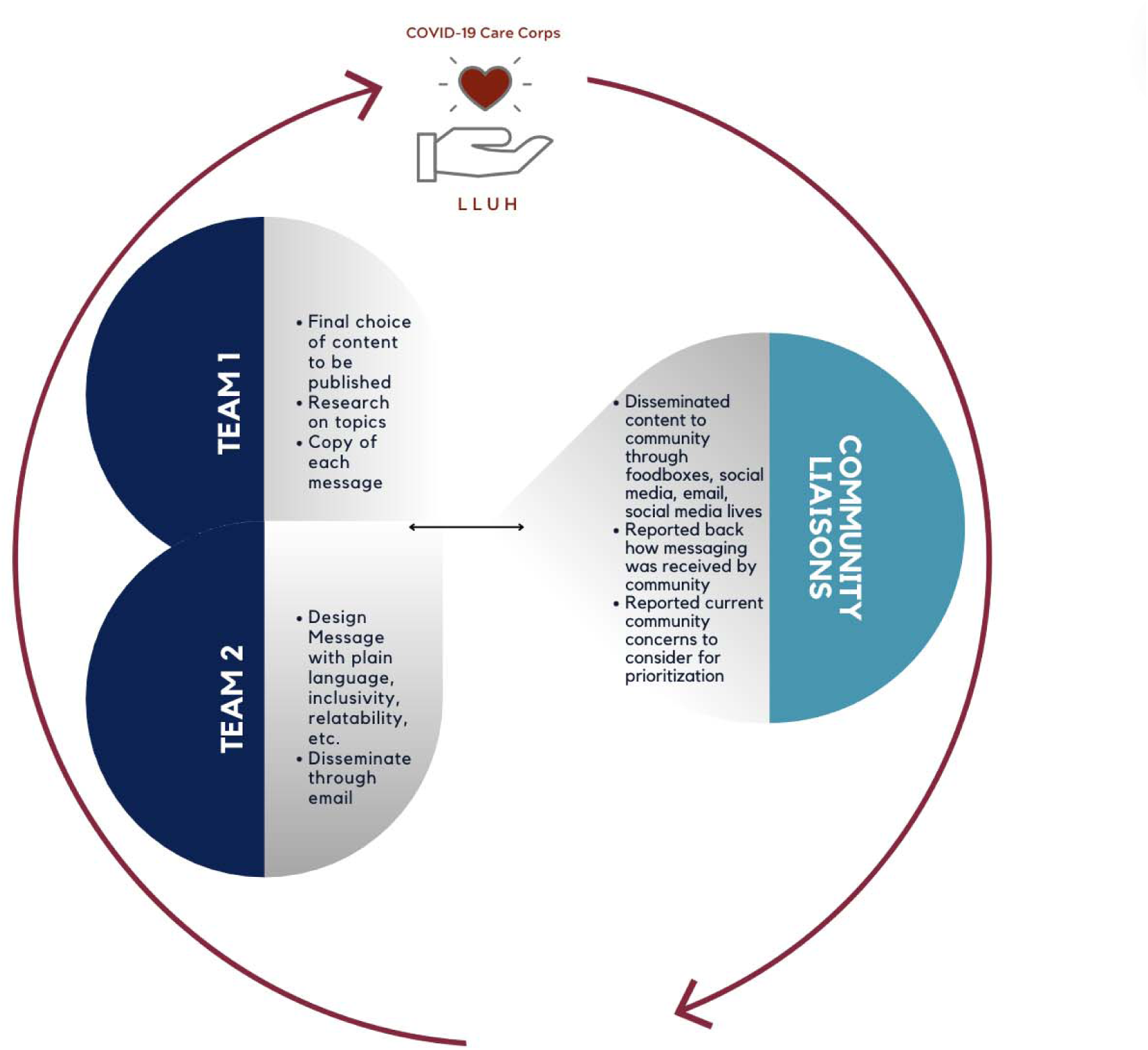
CCC Structure and Process. Note: teams from Loma Linda University and community liaisons met on a bi-weekly basis to discuss community concern to consider in new messaging as well as feedback and response from previous messaging to address future dissemination methods and design.

Team two collaborated with community partners to disseminate information through the appropriate channels in a timely manner and included the liaisons. When content was ready to be shared, it was sent out to liaisons via email and also uploaded to a website that archived all the content created. Liaisons then passed on the content through email blasts, sharing on social media accounts or printing handouts. Organizations that provided footboxes included handouts with every box for maximum reach.

Both teams came together every week to decide on what topics to include for the next week’s content development. This was informed by liaisons sharing what their populations were asking about and specific mandates that were directly impacted by. The content, for example, mask mandates were in place, but accessing masks was difficult for some. Then content was developed sharing not only where individuals could find free masks but why masks were important to mitigate spread. So liaisons were the key component in closing the feedback loop by disseminating information but also informing the team to guide future health information.

### Data and Analysis

Because of the emergency state of the pandemic brought and the need for immediate communication, community didn’t have a direct voice in the feedback. Getting community feedback would have taken time to implement and been difficult to get, since trying to help on a project like this was the last thing on community’s mind when trying to survive. In addition, they were already talking to the liaisons that were chosen for the project and sharing their concerns and needs. They also already had an established trust with the liaisons making the feedback they shared with them more valuable than a formal solicitation. In order to evaluate that process a content analysis was conducted of meeting notes and the measure of interest was how the topics shifted after implementing the feedback loop. However, we noted the result of the emails, i.e. the topics and content that were included in the educational and informational content and ultimately reached the communities as a direct result of this engagement. Thus, for the purposes of this study we considered the outcome of content a proxy for the feedback itself reflecting *the real-time concerns of* the community and in turn becoming the source of the primary data for the feedback loop.

Initially, content created by CCC consisted of translations and interpretations of messaging from federal, state and local health offices. After the implementation of the community “feedback loop”, there was a clear shift in the prioritization of content. To document this shift, all content created from the first CCC message to the last CCC message was categorized into five categories: legal, community, health, COVID-19 and mental health. The number of messages within each category was then divided by the total number of email messages sent out to the broader community to calculate the percentage of a topic sent out in each email. This percentage was then charted over time. A marker was placed at the initial implementation of the feedback loop as well as the first time the COVID vaccine was made available. More details about the “feedback loop” process can be found below.

To reflect on the initiative and learn from the processto better implement similar strategies in the future, community liaisons and project managers who were part of the CCC initiative were interviewed. Six key informant interviews were conducted over Zoom using a semi-structured guide to inquire about the benefits of the program, priorities, how content was shifted, select aspects of community collaboration and the process of incorporating community voice.

Participants gave verbal consent to participate in the interview and be recorded before the interview and recording started. Interviews lasted between 20-40 minutes, were audio recorded and then transcribed using Trint software. After transcripts were verified, each transcript was then coded by the same coder using inductive emergent, high-level coding. The codes were then organized hierarchically, and recurring themes were recorded by a different coder.

## Results

### Feedback Loop: Content Prioritization Results

Illustrated in Figure 2 are the changes in content prioritization after the feedback loop was incorporated. On the x-axis is a timeline from the formation of CCC to approximately a year later when the direction shifted and CCC was absorbed into other projects. Created content (percentage of messages that included a given topic) is depicted on the y axis by.

**Figure 2.**
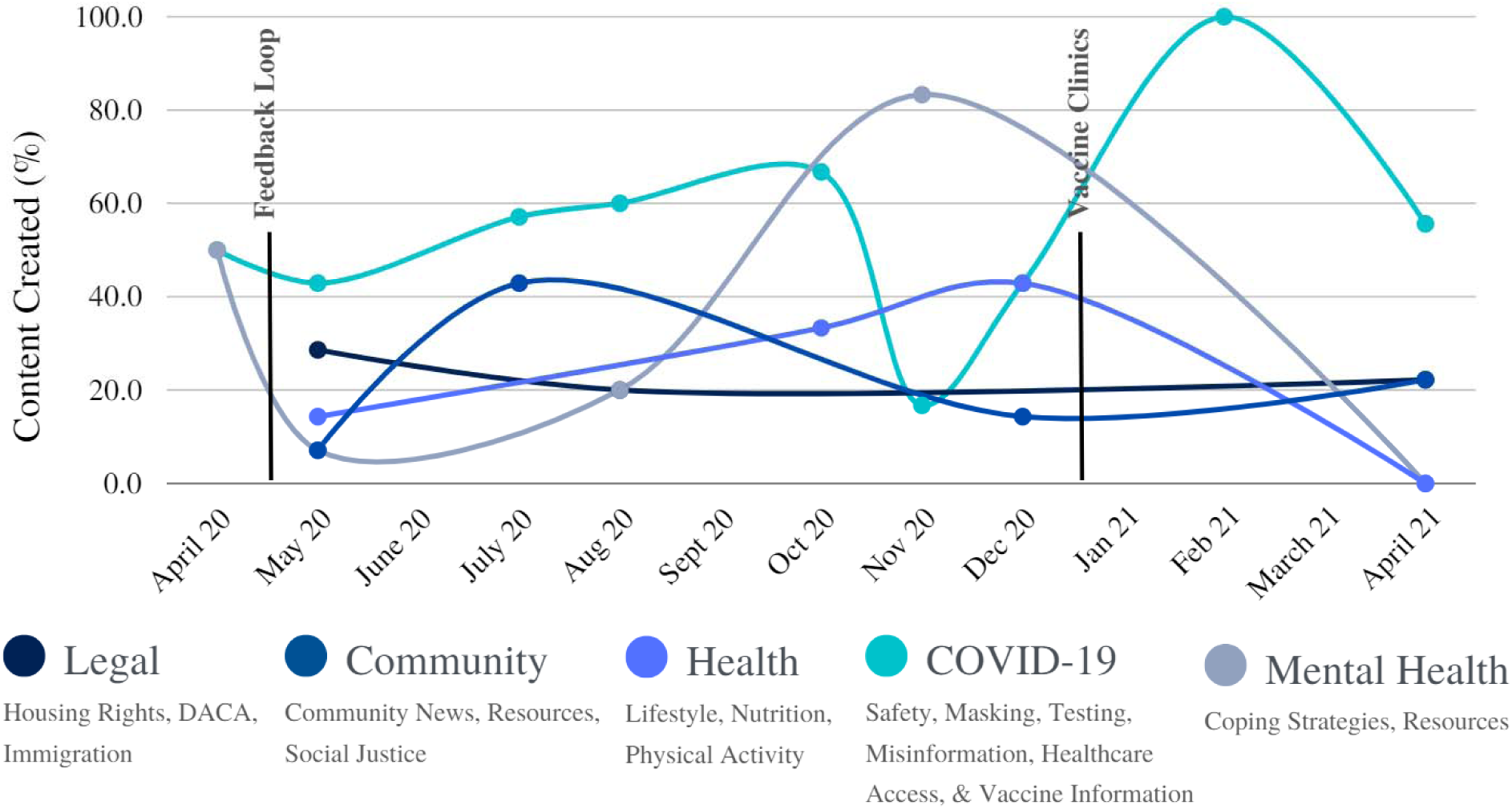
Depiction of informational content priority shifts during the first year of the pandemic.

Each point on the graph represents the intersection between the month a topic was included in an email and the percentage of messages that included that topic. The two markers indicate the first meeting with the community asking about the reception of the content shared up to that point, and the opening of the first vaccine clinic in the area. Although messaging was intended to be shared monthly, there were months associated with the certain periods on the academic calendar (i.e. summer break, new academic year) where messaging was not sent out

Consistent messaging regarding Covid-19 was included in every email and increased with the availability of the vaccine. Yet, there was a clear need for other health and social information, topics regarding immigration and housing rights, lifestyle, nutrition, and community news and resources. Specific topics coincided with current events like Immigration and Customs Enforcement (ICE) raids, evictions, racial tensions, inflation causing an increase in grocery prices and other social and environmental issues.

### Liaison KII Results

Two broad themes emerged from the key informant interviews that capture the essence of Covid Care Corps: information and community voice (see Table 2). Both themes arose as major issues to address during the COVID-19 pandemic with unreliable information as a barrier and the incorporation of community voice as a solution.

**Table 1.** Quotes related to prominent themes from KII Interviews.

**Figure 3. COVID-19 Pandemic Timeline and Loma Linda University (LLU) Response.**
| Table 1. Quotes related to prominent themes from KII Interviews |  |
| --- | --- |
| Themes | Quotes |
| **Information** |  |
| Access | *"...have these fliers, you know, reminding them, you know, that step by step, how do they can, you know, like, prevent...People receive that information."*<br>*"I think it was definitely in alignment with what the community like, the questions that we had. So it was a reasonable response to that and also any needs. And then I think it was it was definitely in alignment."* |
| Health Literacy | *"...because I think a lot of times, if you're someone who hasn't had the opportunity to, you know, get higher education, especially in this country, this language, etc., then you see information and you kind of take it at face value. And you may not have the tools or skills that are taught to be able to kind of pass through it and, you know, read an entire news article or read an entire like, journal article, like you don't even know how to, like, navigate those things, especially if it's not in your first language."*<br>*"There's a lot of various different forms of information that were coming out. You know, like you said, we had a president that was probably not doing the best job as a president in general, but also, guiding us through a pandemic. And so that was very difficult for us to get a lot of backlash and to prove to people that, you know, there's science behind what we're trying to tell them."* |
| Digital Literacy | *"The need of information from credible sources was definitely a big need. And where to access those services, especially since a lot* |
|  | *of things were being found online”*<br>*“...to provide information to community members was really important. And, I feel like by breaking that down right through the content that was available, I think it really served, an important role”*<br>*“It was also like how to navigate different platforms, even how to navigate like, text messages or, you know, information on what, like, one's own phone was really key.”* |
| **Community Voice** |  |
| Collaboration | *“I think it's always great to like, you're part of those spaces because I also hear like feedback from what's happening in other communities or like content was raised like, oh, like, you know, we need this over here.”*<br>*“It's it's just a great example of how people can come together to respond to an emergency and even a non emergency, like, I think it's really one of those spaces that's really collaborative right from the beginning and had just a lot of alignment in like, we're here to respond to community needs, as soon as possible. And, I really appreciated that framework that we worked, you know, with. And, it'd be amazing to just see it replicated and built upon.”*<br>*“I didn't really get to partner with folks in other counties or let alone outside of the eastern Coachella Valley because of like, just, we're like kind of like distance far away from things and things like that. So I know just a highlight was just having that access and to connect with other folks.”* |
| Trust | *“To rely on that, they could say, ‘well Loma Linda is in my backyard, so I can at least rely on what they're saying’ is true. And I think to even more. More so that they could connect faces to that, well masked faces right here. These are these are the people who are writing that message.*<br>*“So there has to be that trust in that relationship, to be able to say, like, hey, like this information that you got, like it's actually like not verifiable, etc...”*<br>*“It's like, are we building that trustworthiness? Is it coming from us? Are we also doing the pieces that put us on the line as well?... any real relationship has to be mutual. It has to be reciprocal. And if it's not, then then we're not building ourselves to be trustworthy and then blaming people for not trusting us.”*<br>*“I was hearing it from so many people in my own circle when we did, you know, vaccine clinics, and they were like, ‘I'm only here because I trust Loma Linda,’ or ‘I'm only here because I trust you,’ or ‘I'm only here because someone told me about it.’ And there's... and I think just that level of trust can only happen when relationships are real”* |
| Cultural Humility & Relevance | *“...speaking Spanish, being able to kind of look at the cultural context of whether the material not only was like correct Spanish, but like, was it something that would make sense culturally to the community in that sense?”*<br>*“Most of the information coming out was in English, which is fine. But was it? It was very academic and most of it was like study and* |
|  | *research in English. And so it's like, 'how do you actually get that translated to the community, not only language wise, right?'*<br>*So we would review the fliers and the information that we made sure it was culturally appropriate to deliver to the community and understandable to make sure that we weren't we're using big words that probably weren't going to be understood.*<br>*"...remembering that everyone is, everyone has important knowledge. This is just the one that we're holding at this time. So being able to learn from people as well and be open to it really being like a learning cycle rather than a one sided, you know, information sharing is really important. And I think, okay, those are relationship pieces. It's what makes that possible, rather than having it just be like, okay, great. Like we're churning out more things, right? Hearing from people that it's actually impacting their lives was the best part. So being able to know that, takes people willing to talk to you, which takes trust."* |

Challenges with information - whether the absence of it, its accessibility, credibility, or the ability to find and evaluate it - were concerns for all participants. This made it difficult for the community to decipher what information to trust and whose information, credible or not, they were likely to follow. Within the theme of information, three sub themes emerged: access, health literacy, and digital literacy. Equally problematic was the source of information and whether that information was credible. Religious groups, family members, and media platforms, such as Facebook, played a significant role as sources of information in the study population’s native languages. Communities also relied on word-of-mouth from those they trusted.

Health literacy also emerged as a critical issue for community members. The pandemic shut down many social gatherings leaving individuals to rely on gathering information through digital sources or their proximal social circles. This posed a challenge for the community as people wondered where to get and evaluate credible information. Information could be found digitally, but navigating fact and fiction required both trust and knowledge. Finding information that was relatable and in a preferred language was yet another challenge.

Critical to accessing information were the basic skills necessary to find, understand and use information to make informed decisions. Digital literacy posed a challenge for the predominantly Spanish speaking community on how to navigate various platforms such as scheduling an appointment with the doctor. Participants noted that they were part of a community with low technical skills and minimal knowledge of where to find evidence-based information about COVID-19 online.

Both a function and a necessity for CCC to address issues within the theme of information was the incorporation of community voice as a remedy. From the theme of community voice emerged four sub themes: collaboration, trust, relevance and cultural humility. Collaboration encompassed working in tandem with community members, local organizations, and with interdisciplinary expertise from the academic partner. Space was created within CCC’s feedback loop method for those with a concern for the community to work collectively to address the challenges that stemmed from the pandemic. CCC leveraged resources and networks to reach the local population in high need.

Trust, as mentioned by all participants, was a key factor for CCC with the local community. Without the existing relationship between LLU and the community, there would be no trust in the academic partner disseminating the information via fliers and media. These were disseminated after they were discussed and reviewed by the community. Since all partners were local, CCC had credibility in the community. Trust also included relationships with the community and reciprocity to build trustworthiness. An example mentioned by a participant was the community’s ability to connect the content from CCC to actual persons from the community. Another interviewee recalls community members specifically mentioning they felt safe receiving the vaccine from the academic partner because of its reputation, as well as trusting the information shared from sources they specifically recognized and had worked with prior to the pandemic, such as CHWs that worked within the existing structure of the academic partner’s programs.

Reviewing documents for relevance before dissemination by community members themselves was amongst the priorities of CCC. Special attention was given to developing culturally and linguistically relevant content and “translating” material in “lay” language was key.

Lastly, a noted approach to incorporating community voice that emerged was cultural humility in relating with the local community. By ensuring that CCC was also learning from the community, it positioned itself as a partner rather than a one-sided authority on information. Community voice with collaboration, trust, relevance and cultural humility enabled CCC to address the initial themes and issues presented within the information theme. As community voice was incorporated into the processes of CCC, content shifted to reflect information that the community prioritized enabling CCC to build rapport with the community as a trusted source of information.

## Discussion

When CCC was first established, health information that came from federal, state and local health departments related to the COVID-19 and mental health were prioritized. The initial focus was translating already existing content to be more relatable and accessible to the Inland Empire community. Once the “feedback loop” was incorporated, there was an expansion of topics and original content. These topics included legal information, health and lifestyle tips and local community information about available resources. Once vaccines were available, information focused on how to access the vaccine, vaccine safety, and side effects.

CCC developed relationships through community liaisons with the community. The community trusted the academic institution “enough” which further supported the tireless work of community health workers (CHWs). These CHWs represented the institution in communities by spending time to find and provide the resources needed. Communities also trusted the local community-based organizations that showed up to create emergency response systems for food boxes or diaper distribution and modeled trust for the academic partner. The inclusion of liaisons to achiev bi-directional communication was a novel communication strategy within the community-academic partnership. To the local community, some SDoH challenges were as great as, and in some cases greater than, concerns about COVID-19. These included access to food, housing, residency, and mental health, in addition to avoiding the virus and mitigating its spread. More individuals were not only doing everything in their power to keep themselves and their families from getting the virus, but also trying to meet basic needs for their families, manage existing health conditions and chronic diseases, keep their children from falling behind in school and showing up to work in conditions that may have increased their risk.

Another concurrent study by Celestin that took place in the same region supported the need for the shift in topics. The study found that several participants had lost their jobs, and there were very real fears of losing homes in addition to many facing food insecurity. There was definitely a concern for the virus itself, contracting it and losing loved ones to it, but there were other competing fears that affected everyday life. There was a fear of what the future would be like. There was also a fear of the overall health care system and their experiences in receiving care (Celestin et al., 2024). The fears and worries identified by this paper were the same fears and issues that were shared to the liaisons emphasizing the need for the teams to address in new information being disseminated (See Figure 4).

**Figure 3.**
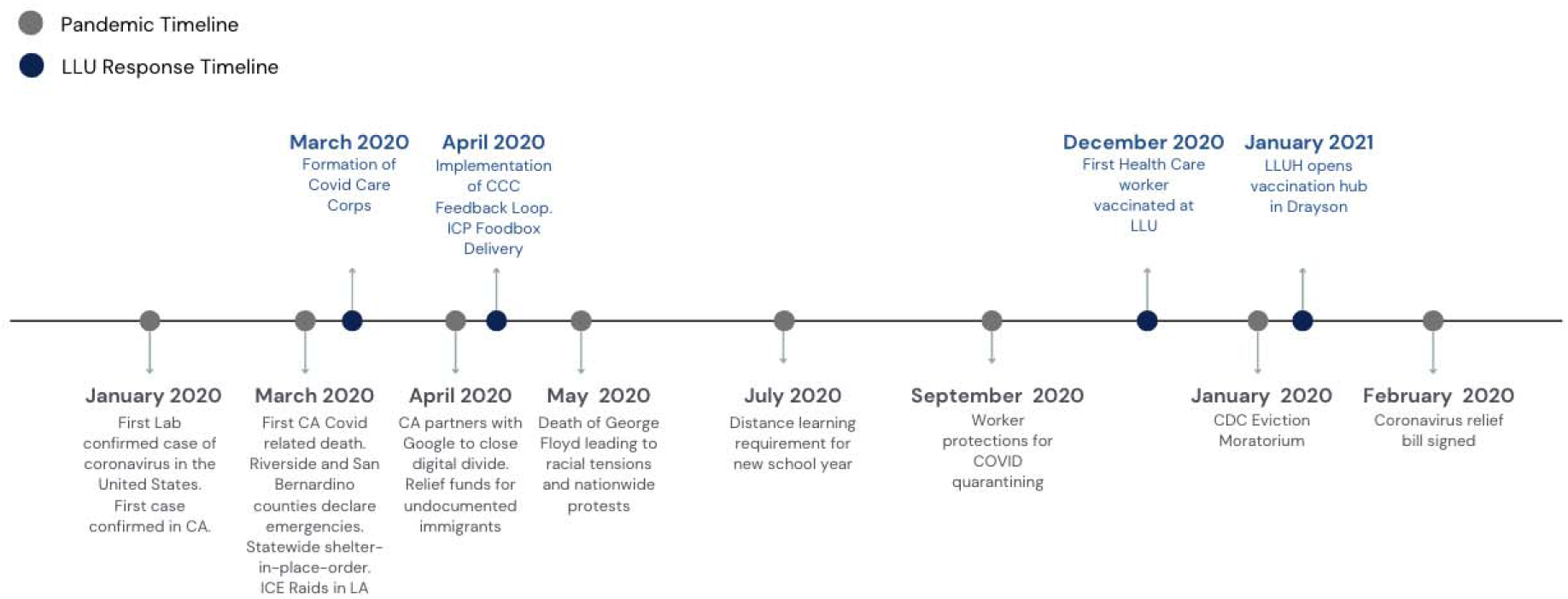
The number of times specific fears were greater than the fear of contracting the COVID-19 infection. COVID-19 Pandemic Timeline and Loma Linda University (LLU) Response.

A topic that was brought to light from the feedback loop was immigration. Individuals were worried about being able to access healthcare and other resources supporting families due to fear of deportation. Rumors about issues with future immigration cases resulting from receiving public assistance may have impacted a family’s decision-making when seeking care or resources. When vaccines were made available, there was a concern around the requirement to show identification and the possible repercussions. On March 19th, 2020, California implemented a stay-at-home order and, the same day, US Immigrations and Customs Enforcement (ICE) officers in Los Angeles raided immigrant communities (Lopez, 2020). The proximity of this event alone impacted approximately 259,000 undocumented residents in the Inland Empire and their willingness to get vaccinated or seek and accept resources (LLUH, 2022).

As shown in the study by Celestin, finances and job security were a major concern. Many feared losing their job, their spouse losing a job, or encountering financial issues (Celestin et al., 2024). Thus, paying bills was a priority. Many of the jobs in the Inland Empire are low wage jobs such as warehouse or transportation jobs and were deemed “essential”, thus increasing risk of exposure (Unidos US, 2021). Many workers were aware of the risk of being exposed to the virus on the job and bringing it home to their multigeneration households, where family members were often at higher risk due to chronic illness or age (Selden & Berdhal, 2021).

Many families in the Inland Empire live in multi-family, multi-generational homes, which made housing insecurity a greater burden, especially with the uncertainty of job security (Vandenberg, 2021). San Bernardino and Riverside counties had some of the highest eviction rates in the U.S. with rates of 42 evictions and 37 evictions per thousand respectively (Herrera & Paluch, 2023). It wasn’t until the COVID-19 Tenant Relief Act that passed in January of 2021, and the CDC Eviction Moratorium in March of that year that there was some relief felt in the region (Proctor, 2021). This is reflected in the secondary data described earlier with a decrease in housing fear as of March of 2021.

At the onset of the pandemic, many were conscious that their overall health affected their risk of contracting the virus. Thus, adequate nutrition became a priority for many families (Lange & Nakamura, 2020). The region is already categorized as a food desert which complicated food access during the pandemic (O’Hara & Toussaint, 2021). Feedback from the community included requests for healthy grocery shopping tips, healthy recipes with food that was available and tips to make food last longer. School lunches and community food box programs did much to alleviate the burden and were a great way of disseminating CCC’S public health information.

All the other concerns mentioned above took a toll on community members. Over 54% of U.S. Americans know someone who passed away due to COVID-19 and approximately 7% of adults had to adjust to living with long COVID (Unidos, 2021). This burden added to the already high mental and emotional toll, due at least in part to the controversial political climate of that time such as the contentious election and racial tensions catalyzed by the death of George Floyd.

Having a constant feedback loop allowed the community to voice and highlight the concerns and issues that they prioritized. Without the bi-directional exchange, the other topics may not have been considered, further disconnecting many from critically essential information. Hearing from diverse voices including community members and local organizations strengthened both the timeliness and relevance of CCC content, but required trust and collaborative relationship.

However, creating this one-time space for community voice and engaging with community is not enough. It is critical to address the concerns and incorporate the knowledge to then implement noticeable, real-time and noticeable changes in the interventions and programs to foster trust, which is what CCC strived for. Through the academic partner, as well as local community-based organizations and entities such as the San Bernardino City Unified School District, CCC provided information about nutrition and vaccinations, but also made food boxes available, and delivering them to families lacking transportation. Partners emphasized the importance of keeping up with school for the youth and helped parents/guardians understand how to use various platforms such as Zoom for work and schoolwork. They also addressed the digital divide by facilitating resources for free internet access in areas where it was needed: schools and local prominent businesses in the Inland Empire provided digital devices free of charge and created hotspots using school buses, to assure internet connectivity (Ekbatani, 2020).

Through this experience, we learned that community voice should not just be a one-time acknowledgement but rather a continuous and integral part of community health program development and evaluation. The community should be a collaborative partner throughout the entire process (Quinn et. al, 2023) as this is vital to ensure the efficacy of the shared health information and will be reflected in the effectiveness of the message (reception and application of that information) within the community.

In addition to feedback on topics, feedback on the content delivery and dissemination was also significant. Content was created for social media and for print but, due to accessibility challenges, it was decided that print would be of priority because dissemination was simple through the food box distribution - an example of the continuous re-evaluation necessary for effective implementation.

The pandemic put everyone in a state of survival creating a unique setting for collaboration to mitigate the spread and burden on health systems and keep everyone healthy. There was no space for internal focus or pre-set political agendas. Equitable, community-centered, bi-directional health communication wa a priority for all allowed the partnership to show that they value the community and its concerns instead of only focusing on their own agendas. For instance, for the academic partner, mitigating the spread and severity of the infection was important, but working with the community to address their concerns allowed the establishment of trust that benefited the overall goal when trying to increase vaccination efforts.

## Strengths and limitations

From the effectiveness of the CCC messaging as shown in the data and the participants’ reflections, it is evident that this project had a positive impact on the local community. The crisis-driven nature of the development of this collaborative left much to trial and error, and unfortunately sustainability was not prioritized. Gaps were left in the process because much of the project was run by students who then graduated, creating poor sustainability. Future implementation of a CCC should continue to include students but with long-term faculty and staff for stability. Another limitation was that community voice was not always directly included and was instead filtered through community liaisons. Future implementation should consider having community members who are not affiliated with any organization on either teams.

## Conclusion

Prioritizing community voice through feedback loops with health communication plans allows for insights into what local community needs are and how to address them, thus facilitating more effective and relevant communication. Academic partners’ priorities may not be the same as those of the community, but taking the time to address community perspectives allows for building pathways of trust. Instead of demanding trust and compliance, creating spaces for collaboration, including community liaisons who already have that trust and leverage and finding common ground can reveal mutual interests. Community engagement in health communication strategies should be a requirement throughout development, implementation and evaluation, rather than an afterthought.

## Data Availability

No data produced in the present study will be available.

## Acknowledgements

The authors are grateful to all the departments and individuals at Loma Linda University who were a part of developing the COVID Care Corps, especially to Crissy Irani and Dr. J.C. Belliard who led the project. Special thanks are also directed to Olivia Vaquera for her work in the community as well as the many other community liaisons and health workers who provided valuable support.

## Funding

The author(s) reported there is no funding associated with the work featured in this article.

## Declaration of Interest

The authors report there are no competing interests to declare.

## Ethical Statement

CCC was a program implemented at Loma Linda University Health and data related was from evaluations of the program. In the study from which the secondary data was collected, key informant interviews and focus groups were performed in accordance with the principles of the Declaration of Helsinki, and ethical approval for the study was obtained from the Loma Linda University Institutional Review Board (IRB #518,068)

## Notes

### Competing Interest Statement

The authors have declared no competing interest.

### Funding Statement

This study did not receive any funding

### Author Declarations

IRB of Loma Linda University gave ethical approval for this work.

